# Prominent role of PM10 but not of circulating inflammation in the link between air pollution and the risk of neurodegenerative disorders

**DOI:** 10.1101/2023.05.17.23289154

**Authors:** Alessandro Gialluisi, Simona Costanzo, Giovanni Veronesi, Assuntina Cembalo, Alfonsina Tirozzi, Stefania Falciglia, Moreno Ricci, Francesco Martone, Gaetano Zazzaro, Marco Mario Ferrario, Francesco Gianfagna, Chiara Cerletti, Maria Benedetta Donati, Stefania Massari, Giovanni de Gaetano, Licia Iacoviello, the Moli-sani Study Investigators

## Abstract

**Background:** Several studies revealed an implication of air pollution in neurodegenerative disorders, although this link and the potential underlying mechanisms remain unclear.

**Objectives:** To analyze the impact of air pollution on neurodegenerative risk by testing multiple pollutants simultaneously, along with other potential risk/protective factors, and the role of circulating inflammation.

**Methods:** In the Moli-sani cohort (N=24,325; ≥35 years; 51.9% women, baseline 2005-2010), we estimated yearly levels of exposure to nitrogen oxides, ozone, particulate matter (PM10), sulfur dioxide and BTX hydrocarbons in 2006-2018, applying residence geo-localization of participants and Kriging interpolation algorithm to land measurements of air pollutants. We performed a principal component (PC) analysis of pollutant levels and tested associations of the resulting PC scores with the incident risk of dementia (AD) and Parkinson’s disease/parkinsonism (PD), through multivariable Cox PH regressions adjusted for age, sex, education level, and several professional and lifestyle exposures. Moreover, we tested whether a composite biomarker of circulating inflammation (INFLA-score) may explain part of these associations.

**Results:** Over 24,308 subjects with pollution data available (51.9% women, mean age 55.8(12.0) years), we extracted three PCs explaining ≥5% of pollution exposure variance: PC1 (38.2%, tagging PM10), PC2 (19.5%, O3/CO/SO2), PC3 (8.5%, NOx/BTX hydrocarbons). Over a median (IQR) follow-up of 11.2(2.0) years, we observed statistically significant associations of PC1 with an increased risk of both AD (HR[CI] = 1.06[1.04-1.08]; 218 cases) and PD (1.05[1.03-1.06]; 405 incident cases), independent on other covariates. These associations were confirmed testing average PM10 levels during follow-up time (25[19-31]% and 19[15-24]% increase of AD and PD risk, per 1 μg/m^3^ of PM10). INFLA-score explained a negligible (<1%) proportion of these associations.

**Discussion:** Air pollution – especially PM10 – is associated with increased neurodegenerative risk in the Italian population, independent on concurring risk factors, suggesting its reduction as a potential public health target.

## Introduction

In the last decades, pollution – prominently air pollution – has represented a hotspot of investigation and received increasing interest from policy makers, in view of the notable burden that it implies for public health and welfare systems [1]. Air pollution is defined as a combination of various compounds originating from different anthropogenic and biogenic emission sources, ranging from particulate matter (PM, with different components based on their aerodynamic diameter) to gases (carbon monoxide, nitrogen dioxide, etc.), and organic chemical compounds (e.g., hydrocarbons) [2]. Not only chronic exposure to air pollution is associated with increased cardiovascular and respiratory diseases risk [3], but recent findings have supported a link between air pollution and neurological disorders, including neurodegenerative disorders like dementia of different origins (hereafter called AD), and Parkinson’s disease or parkinsonisms (hereafter called PD) [5,6]. Dementia is the most prevalent neurodegenerative disorder worldwide, is characterized by memory loss and cognitive impairment and may be of different types, the most frequent being vascular dementia and Alzheimer’s disease. The latter represents 60-80% of all dementia forms [7] and is characterized by typical accumulations of amyloid beta and tau proteins aggregates in the brain, leading to neuronal loss in cortical areas and brain atrophy. Parkinson’s disease represents instead the second most common neurodegenerative disease, characterized by alpha-synuclein aggregates accumulation in dopaminergic neurons of the substantia nigra, which leads to progressive, irreversible loss of these neurons. This manifests through muscular rigidity, bradykinesia, tremor of resting limbs, gait and balance impairments, and often to a progressive loss of cognitive functions [8]. Moreover, Parkinson’s disease often shows several common characteristics with Parkinsonisms - clinical syndromes with similar manifestations, although of different origins, like vascular parkinsonism and multiple system atrophy [9,10]. Interestingly, both AD and PD are often comorbid, show a partial overlap of signs and symptoms and are triggered at least in part by the same molecular mechanisms, like circulating and neuro-inflammation [11,12].

While recent reviews and meta-analyses have reported growing evidence suggesting that chronic exposure to air pollution – especially to particulate matter with aerodynamic diameter < 2.5 μm (PM2.5) and nitric dioxide (NO2) – is associated with an increased risk of both incident and prevalent neurodegenerative disorders like AD and PD [13,14,15,16], these associations are often not replicated and only partly concordant across the different studies, for several reasons. Among them, the heterogeneity of study settings probably represents one of the main hindrances to results concordance, with very scarce longitudinal studies carried out in large real cohorts (e.g. [17]) and many administrative cohort (e.g. [18,19,20]), case-control studies (e.g. [21,22]) and geostatistical analyses (e.g. [23]) being reported. Indeed, most of these studies tested a handful of pollutants simultaneously, but only few analyzed them together with other potential factors influencing neurodegenerative risk. As a consequence, these studies often suffer from residual confounding bias due to scarce adjustment for other potential confounders of the association between air pollution and neurodegenerative risk, like lifestyles and professional covariates. Other studies are mostly based on a case-control approach – due to the relatively low prevalence of these disorders – and hence may suffer from reverse causality bias. Overall, very few studies are based on deeply characterized longitudinal cohorts (e.g. [17]) which may allow to disentangle the cluster of risk/protective factors for neurodegenerative disorders, including sociodemographic, lifestyles and professional factors.

The main aim of this work was to test how air pollution may influence neurodegenerative risk – in particular the incident risk of the most prevalent neurodegenerative disorders like dementia and Parkinson’s disease/parkinsonisms – independent on other potential risk/protective factors. We did this in a general Italian population cohort, with a deep phenotypic assessment, by simultaneously analyzing the influence of several air pollutants, sociodemographic, lifestyle and professional exposures, over a 12 years follow-up. Thanks to the availability of molecular blood markers, we also tested a potential role of circulating inflammation in the links identified, looking for further support to previous evidence reported in animal models [6,24,25].

## Subjects and Methods

### Population of study

The study population consisted of participants to the Moli-sani project (N = 24,325; 51.9% women), a cohort of Italian residents recruited from the general population of Molise Region (Central-Southern Italy), between March 2005 and April 2010. Exclusion criteria were pregnancy at the time of recruitment, inability to understand terms of participation, current poly-traumas (i.e., simultaneous injury to several organs or body systems), coma, or refusal to sign the informed consent [26]. The Moli-sani Study was approved by the ethical committee of the Catholic University of Rome (approval nr: P99, A-931/03-138-04/CE/2004, 11 February 2004) and all the participants provided written informed consent.

### Geolocalization

For this study, participants were geolocalized based on their residence address – available for all the participants - with the use of complementary dedicated software APIs, like Geokettle, QGIS and Here (see URLs). The data were cleaned up manually correcting the errors detected, such as incorrect postal codes. To facilitate the geocoding process, the addresses were transformed by replacing the abbreviations. Using an automated procedure based on the Here API, it was possible to assign latitude and longitude coordinates for all the subjects in the database. A handful of subjects with residence outside the Molise region (n=17) were geolocated out of the Molise region and hence removed from the analysis. The results of the geocoding were verified using the shapefiles of the boundaries of the municipalities of the region. Addresses placed outside the limits of the city of residence were manually geolocated using Google Maps. With this procedure, it was possible to assign latitude and longitude coordinates for 24,308 subjects in the database with a high level of confidence. The subjects were then linked to air pollution maps (built as described below), which allowed us to estimate the amount of exposure to each pollutant in the physical coordinates where subjects reported their residence.

### Air pollution exposure

We estimated yearly levels of exposure to ten different pollutants, including Nitrogen oxides (NOX, NO, NO2), ozone (O3), particulate matter with aerodynamic diameter < 10 μm (PM10), Sulfur dioxide (SO2), Carbon monoxide (CO) and BTX hydrocarbons (benzene, toluene and xylene) in 2006-2018. To this end, we applied ordinary Kriging interpolation algorithm to land measurements of air pollutants made publicly available from the regional environmental authority (Agenzia Regionale per la Protezione Ambientale del Molise - ARPA Molise, see URLs). Indeed, Kriging algorithm allows geostatistical data interpolation based on available land measures, to infer also unsampled points across the spatial field [27] (see URLs). To apply geostatistical algorithms and display analysis results on maps, we used ESRI ArcGIS tool, a proprietary geographic information system (GIS; see URLs) which allowed us to display air monitoring stations on the regional Molise map and finally build maps of exposure to different air pollutants (see Figure 1). This algorithm was applied to each pollutant for each year of the follow-up time considered, over participants with physical coordinates available within the Molise region. This returned 24,308 subjects with environmental exposures (each with ten pollutants levels × 13 years). Since Kriging returned exposure intervals which were largely overlapping but with variable limits across the years, to reduce collinearity among variables, we decided to apply a principal component analysis (PCA), so to derive latent variables which could capture most of the shared variance across the 130 variables.

**Figure 1.**
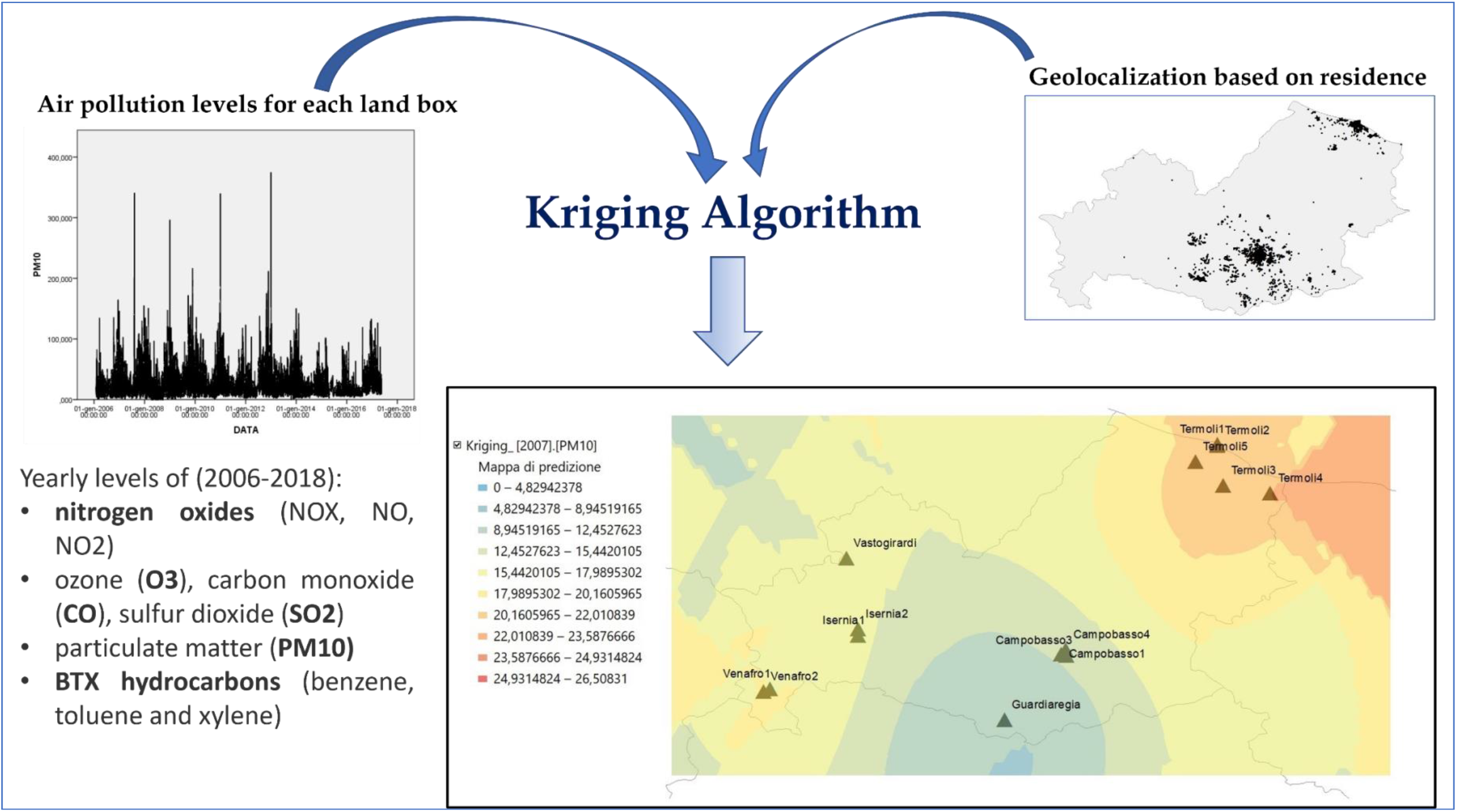
Model used to build air pollutants exposure maps in the Moli-sani cohort.

To this end, we removed toluene levels for year 2017 – which presented a single level of exposure. Then, we computed the mean of the two limits for each level of exposure (or rank) and each pollutant, so to have point estimates of exposure for applying the PCA. We carried out a preliminary Kaiser-Meyer-Olkin (KMO) test of sampling adequacy, which revealed a very good factorability (MSA = 0.97), and a Bartlett’s Test of Sphericity, which suggested a significant discrepancy with the identity matrix, hence notable reciprocal correlations across the variables, which further supported our choice. PCA was performed through Singular Value Decomposition with orthogonal (varimax) rotation (built-in *prcomp()* function in R[28]). The resulting PC scores were then tested as main exposures in survival analyses, as detailed below.

### Outcome: incident neurodegenerative risk

Definition of incident neurodegenerative disorders was carried out through linkage with Electronic Health Records (EHRs) databases like the Italian National mortality (ReNCaM) registry, the Molise regional registry of hospital discharge records (HDRs) and the regional drug prescription registry, using fiscal code of each participant as unique identifier.

Specifically, Alzheimer’s Disease/Dementia (AD) and Parkinson’s Disease/Parkinsonisms (PD) were defined as the occurrence of at least one of the following events up to December 31^st^ 2018 (Table S1): i) Parkinson’s and Alzheimer’s disease reported as main cause of death (ICD-9 cm codes 332 and 331) in the ReNCaM registry; ii) Parkinson’s disease or any parkinsonism, Alzheimer’s disease or other dementias reported as main cause of hospitalization in HDRs (ICD-9 cm codes: 332, 332.0 and 332.1 for PD and 331, 331.X for AD); iii) specific drug prescriptions for the treatment of these disorders in the regional drug prescription registry, like antiparkinsonian (ATC code: N04) and antidementia drugs (N06D).

### Statistical analysis

All statistical analyses of the present manuscript were carried out in R (see URLs)[28]. After removing prevalent AD (n=16) and PD (n=52) cases, as well as those participants with missing information on incident events (0 for AD and 236 for PD), we tested incident AD/PD risk vs air pollution exposure through multivariable Cox Proportional Hazards (PH) regression models, implemented through the *coxph()* and the *surv()* functions of the survival package [29]. To test potential influences of air pollution independent on other clusters of risk/protective factors, multivariable models were built, incrementally adjusted for i) age, sex and education level completed (baseline, Model 1); ii) professional factors like exposure to toxic compounds and professional working class (Model 2); and iii) lifestyles (smoking and drinking status, adherence to Mediterranean Diet, physical activity in leisure time) or their proxies, e.g. body mass index (BMI). A detailed description of the covariates is reported in Supplementary Materials. The main pollution exposure was initially tested as PC scores, as computed through the PCA over all yearly pollutant levels (see above), which were tested all together in multivariable models. If any PC score showed a significant association, levels of the single pollutant/s tagged by that PC were tested, averaged over all the actual years of follow-up for each participant. Sensitivity analyses were carried out removing all early onset cases (<50 years for PD, <65 years for AD), which are more likely to be mostly of genetic origins. All the models showed Variance Inflation Factors (VIF) < 2, suggesting negligible collinearity bias. A Bonferroni correction for multiple testing of three environmental principal components and two main neurodegenerative outcomes was applied, resulting in a corrected significance threshold α = 0.008.

### Testing the role of circulating inflammation

We tested circulating inflammation as a variable possibly explaining the association between air pollution and neurodegenerative risk, as hypothesized elsewhere [8,13,30,31]. Specifically, we used a composite blood-based inflammation index, called INFLA-score, based on four circulating biomarkers - C-reactive protein levels (CRP), blood platelet count (Plt), white blood cell count (WBC), and granulocyte-to-lymphocyte ratio (GLR) – and capturing both serum and cellular-circulating inflammation [32]. This score has been already validated as a comprehensive index of circulating inflammation, since it includes cytokine-related, hemostatic and immune components of the inflammatory response [33], and has been previously associated with the inflammatory potential of diet within the Molisani study [34].

For all potential explanatory markers, the analysis was carried out through bootstrap-based Cox PH regression models (over 1,000 bootstraps). For each bootstrap sampling, we calculated the change in log (hazard ratio) between the model without and with the investigated marker, divided by the log (hazard ratio) of the model without the investigated marker. Then, a proportion of total effect explained (PTE) by the marker was computed as the absolute value of this estimate, and standard deviation (SD) and p-values were defined using the empirical distribution of coefficients resulting from all bootstraps.

## Results

Table 1 reports main sociodemographic and epidemiological characteristics of the analyzed samples both for AD and PD incident risk. Compared to participants of the Moli-sani cohort excluded from analyses based on the lack of concordant geolocalization, lack of follow-up information or prevalent AD/PD condition, analyzed subjects showed a generally higher education level (p = 0.002 for AD and < 0.0001 for PD analysis) and a lower prevalence of CVD, cancer, diabetes, hypertension (p < 0.0001 for both AD and PD) and hyperlipidemia (p = 0.007 for AD, p < 0.0001 for PD), a lower frequency of moderate drinkers and a higher frequency of heavy/very heavy drinkers (p < 0.0001 for both AD and PD). No significant difference in sex distribution was observed (48.11% men in the analysis of AD and 48.15% in the analysis of PD), as well as in all the variables tagging lifestyles, except for leisure time physical activity, which was higher in the analyzed compared to the removed samples (p = 0.03 for AD and p < 0.0001 for PD), and for BMI, which was slightly lower in the analyzed sample of the PD analysis (p = 0.03). No significant difference was found in the distribution of inflammatory markers, except for a marginally lower GLR value in the subjects analyzed for PD, compared to those removed (Table 1).

**Table 1.**
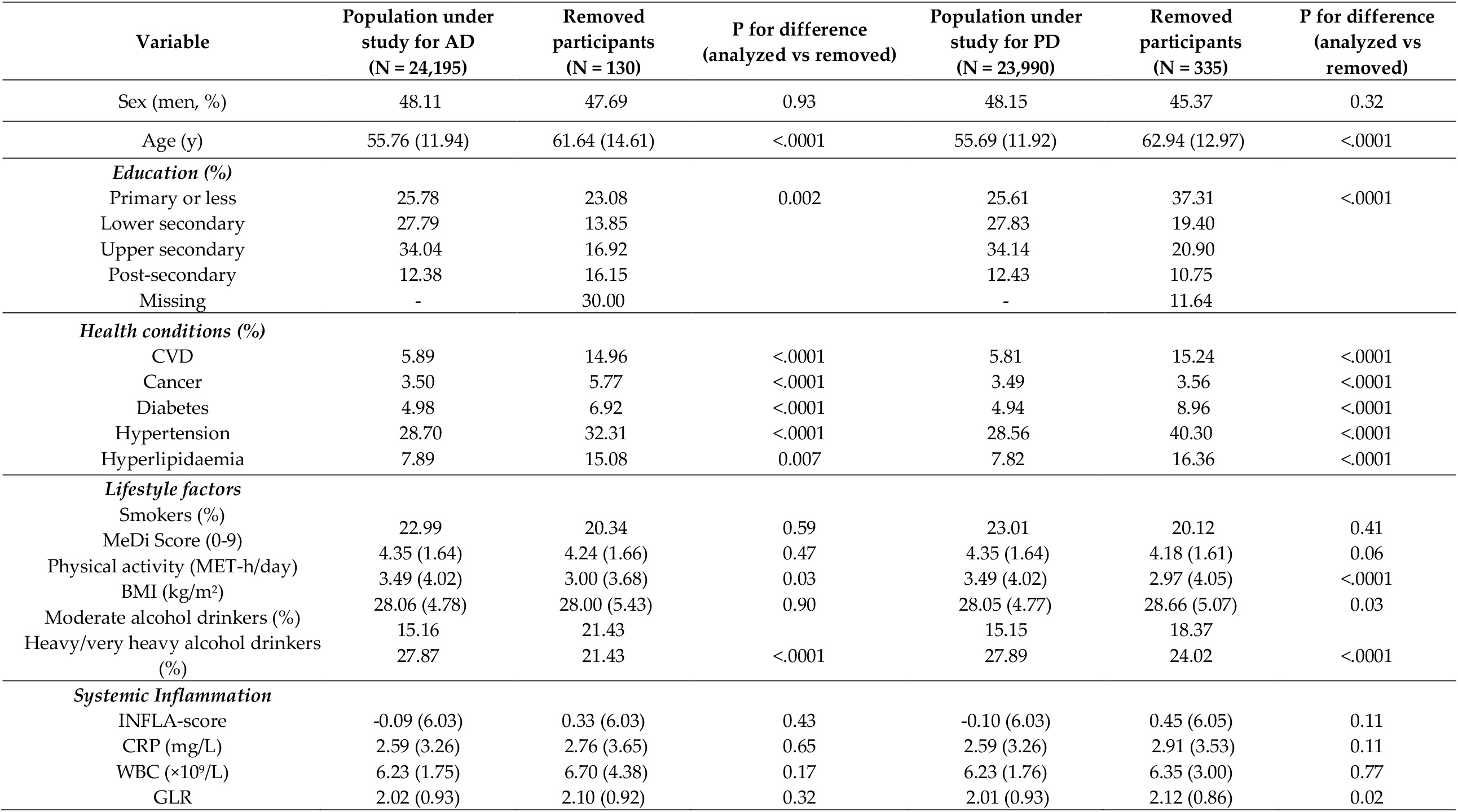
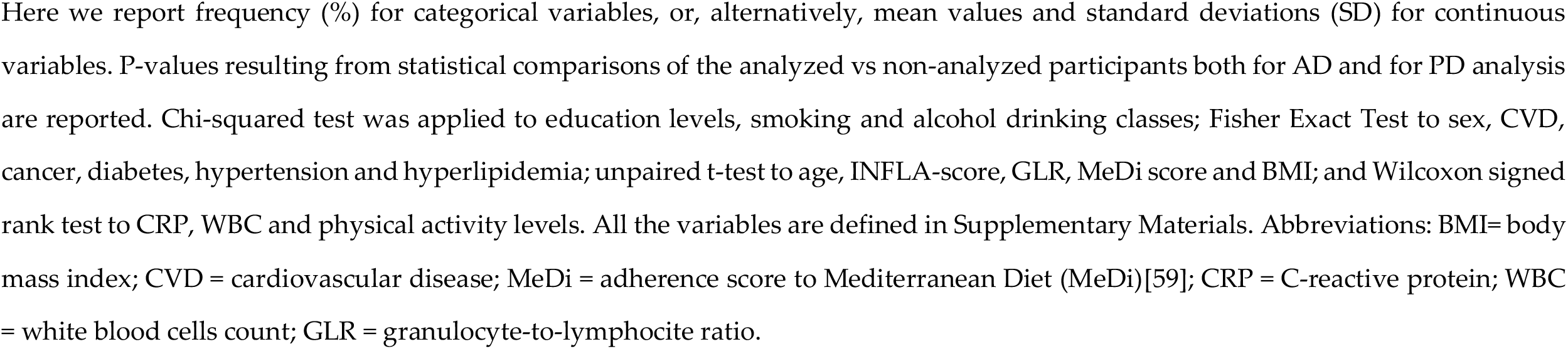
Baseline characteristics of the population under study.

| Variable | Population under study for AD<br>(N = 24,195) | Removed participants<br>(N = 130) | P for difference<br>(analyzed vs removed) | Population under study for PD<br>(N = 23,990) | Removed participants<br>(N = 335) | P for difference<br>(analyzed vs removed) |
| --- | --- | --- | --- | --- | --- | --- |
| Sex (men, %) | 48.11 | 47.69 | 0.93 | 48.15 | 45.37 | 0.32 |
| Age (y) | 55.76 (11.94) | 61.64 (14.61) | <.0001 | 55.69 (11.92) | 62.94 (12.97) | <.0001 |
| *Education (%)* |  |  |  |  |  |  |
| Primary or less | 25.78 | 23.08 | 0.002 | 25.61 | 37.31 | <.0001 |
| Lower secondary | 27.79 | 13.85 |  | 27.83 | 19.40 |  |
| Upper secondary | 34.04 | 16.92 |  | 34.14 | 20.90 |  |
| Post-secondary | 12.38 | 16.15 |  | 12.43 | 10.75 |  |
| Missing | - | 30.00 |  | - | 11.64 |  |
| *Health conditions (%)* |  |  |  |  |  |  |
| CVD | 5.89 | 14.96 | <.0001 | 5.81 | 15.24 | <.0001 |
| Cancer | 3.50 | 5.77 | <.0001 | 3.49 | 3.56 | <.0001 |
| Diabetes | 4.98 | 6.92 | <.0001 | 4.94 | 8.96 | <.0001 |
| Hypertension | 28.70 | 32.31 | <.0001 | 28.56 | 40.30 | <.0001 |
| Hyperlipidaemia | 7.89 | 15.08 | 0.007 | 7.82 | 16.36 | <.0001 |
| *Lifestyle factors* |  |  |  |  |  |  |
| Smokers (%) | 22.99 | 20.34 | 0.59 | 23.01 | 20.12 | 0.41 |
| MeDi Score (0-9) | 4.35 (1.64) | 4.24 (1.66) | 0.47 | 4.35 (1.64) | 4.18 (1.61) | 0.06 |
| Physical activity (MET-h/day) | 3.49 (4.02) | 3.00 (3.68) | 0.03 | 3.49 (4.02) | 2.97 (4.05) | <.0001 |
| BMI (kg/m<sup>2</sup>) | 28.06 (4.78) | 28.00 (5.43) | 0.90 | 28.05 (4.77) | 28.66 (5.07) | 0.03 |
| Moderate alcohol drinkers (%) | 15.16 | 21.43 |  | 15.15 | 18.37 |  |
| Heavy/very heavy alcohol drinkers (%) | 27.87 | 21.43 | <.0001 | 27.89 | 24.02 | <.0001 |
| *Systemic Inflammation* |  |  |  |  |  |  |
| INFLA-score | -0.09 (6.03) | 0.33 (6.03) | 0.43 | -0.10 (6.03) | 0.45 (6.05) | 0.11 |
| CRP (mg/L) | 2.59 (3.26) | 2.76 (3.65) | 0.65 | 2.59 (3.26) | 2.91 (3.53) | 0.11 |
| WBC (×10<sup>9</sup>/L) | 6.23 (1.75) | 6.70 (4.38) | 0.17 | 6.23 (1.76) | 6.35 (3.00) | 0.77 |
| GLR | 2.02 (0.93) | 2.10 (0.92) | 0.32 | 2.01 (0.93) | 2.12 (0.86) | 0.02 |

Here we report frequency (%) for categorical variables, or, alternatively, mean values and standard deviations (SD) for continuous variables. P-values resulting from statistical comparisons of the analyzed vs non-analyzed participants both for AD and for PD analysis are reported. Chi-squared test was applied to education levels, smoking and alcohol drinking classes; Fisher Exact Test to sex, CVD, cancer, diabetes, hypertension and hyperlipidemia; unpaired t-test to age, INFLA-score, GLR, MeDi score and BMI; and Wilcoxon signed rank test to CRP, WBC and physical activity levels. All the variables are defined in Supplementary Materials. Abbreviations: BMI= body mass index; CVD = cardiovascular disease; MeDi = adherence score to Mediterranean Diet (MeDi)[59]; CRP = C-reactive protein; WBC = white blood cells count; GLR = granulocyte-to-lymphocyte ratio.

**Table 2.**
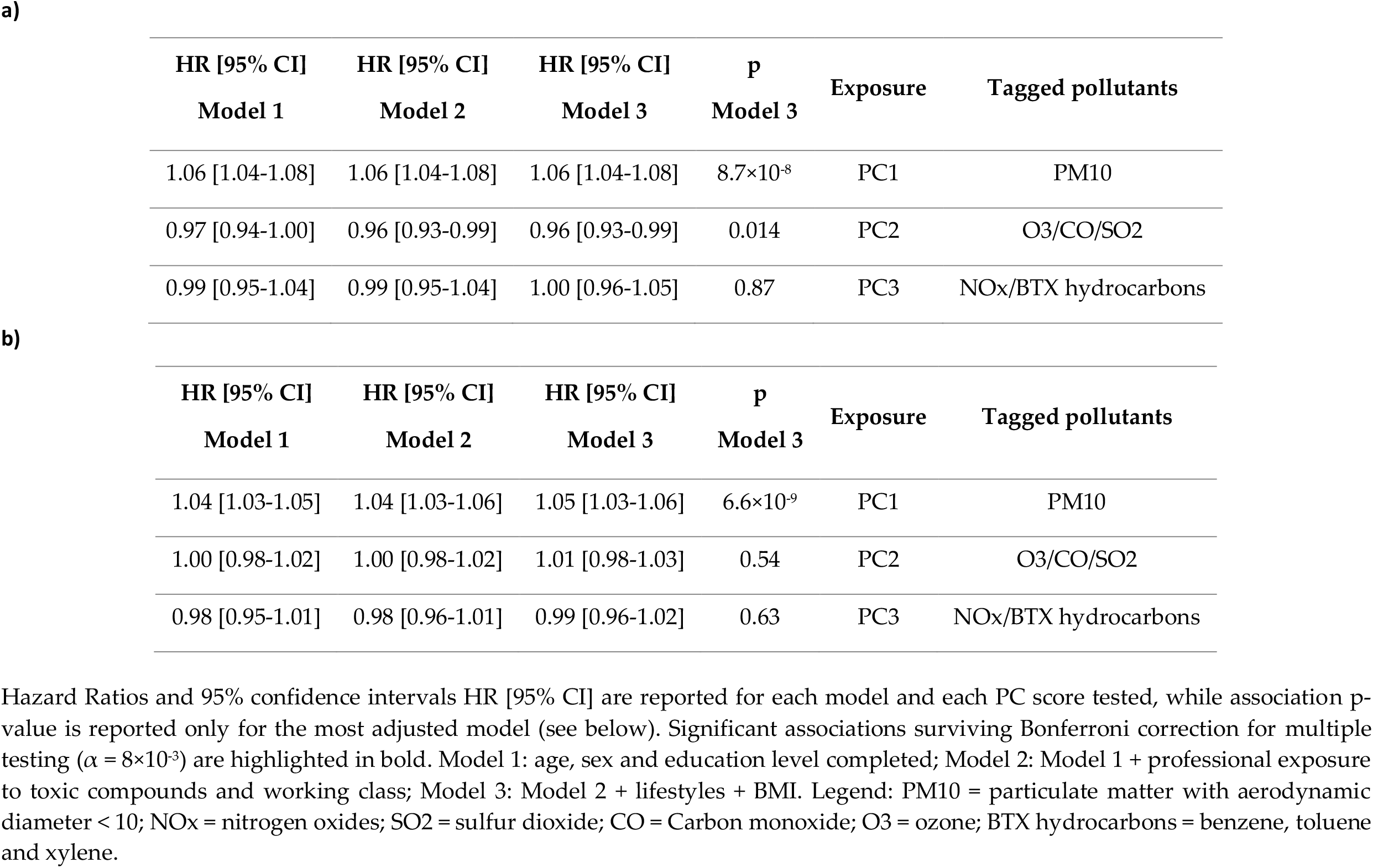
Results of the main analysis of incident a) AD and b) PD risk vs principal components of air pollutants exposure.

A PCA over 24,308 participants with air pollution levels available within the Molise region revealed three main principal components scores, explaining >5% of common variance across all environmental variables tested: PC1 (38.2%), PC2 (19.5%) and PC3 (8.5%; Figure S1). While PC1 was quite clearly tagging PM10 levels, PC2 and PC3 showed moderate to high loadings of O3/CO/SO2 and NOx/BTX hydrocarbons, although less clearly (Figure S2a, b, c). This pattern was consistent with the Spearman’s correlation patterns across all the pollutants tested together, averaged across the years (Figure S3).

Although Model 3 (adjusted for sociodemographic, professional and lifestyle factors) was considered as the main model of reference for the interpretation of results, we report below the results of all the incrementally adjusted Cox PH models to appreciate the influence of incremental adjustment on the associations analyzed (Table 3). Over 24,195 subjects analyzed for AD risk (218 incident cases, median (IQR) follow-up 11.17 (2.02) years), we observed a significant association of PC1 with an increased risk of dementia (HR [95% CI] = 1.06 [1.04-1.08] per unitary increase of PC1 score), which was stable across models adjusted for professional (1.06 [1.04-1.08]) and lifestyles covariates (1.06 [1.04-1.08]; p-value < 8.7×10^−8^). A significant association was also observed for incident Parkinson’s disease/parkinsonisms risk (N = 23,990, 405 incident cases, median (IQR) follow-up: 11.17 (2.03) years), although with a slightly smaller effect size (1.04 [1.03-1.05]), which was again confirmed after adjustment for professional covariates and became even larger after adjusting for lifestyles or their proxies (1.05 [1.03-1.06]; p = 6.6×10^−9^). No other PC score showed significant associations surviving Bonferroni correction for multiple testing, with any of the disorders tested (Table 3), nor any other covariate used in the model, except for age (Table S2a, b). Since PC1 showed high loadings of PM10 and was therefore clearly tagging the levels of this pollutant, we tested directly PM10 for association with incident AD and PD risk, which revealed relative risks consistent with those observed for PC1. Indeed, each unitary (μg/m^3^) increase of PM10 was associated with a 25 (19-31)% increase of incident AD risk and a 19 (15-24)% increase of PD risk in the most conservative model (Table 3, Model 3). Since PM10 showed a bimodal distribution (Figure S4), to ensure against potential biases resulting from potential departures from normality, we compared participants above and below the median level of PM10 in the analyzed population (11.6 μg/m^3^), which revealed strong associations, in line with the effect sizes observed per unitary increase. Participants exposed to average PM10 concentrations > 11.6 μg/m^3^ showed a ∼22x (13-39) increase of AD risk and a ∼14x (10-21) increase of PD risk, compared to subjects exposed to PM10 concentrations ≤ 11.6 μg/m^3^ (Figure 2, Table 3). Associations remained stable after removal of early onset AD and PD cases (Table S3).

**Table 3.** Results of the main analysis of incident AD and PD risk vs PM10 levels.

| HR [95% CI]<br>Model 1 | HR [95% CI]<br>Model 2 | HR [95% CI]<br>Model 3 | p<br>Model 3 | Outcome | Exposure |
| --- | --- | --- | --- | --- | --- |
| 1.22 [1.18-1.28] | 1.24 [1.18-1.29] | 1.25 [1.19-1.31] | 7.5×10<sup>-20</sup> | AD | PM10 (µg/m<sup>3</sup>) |
| - | - | 22.12 [12.51-39.14] | 1.9×10<sup>-26</sup> | AD | PM10 (high vs low exposure) |
| 1.18 [1.14-1.21] | 1.19 [1.15-1.22] | 1.19 [1.15-1.24] | 1.5×10<sup>-23</sup> | PD | PM10 (µg/m<sup>3</sup>) |
| - | - | 14.24 [9.90-20.47] | 1.5×10<sup>-46</sup> | PD | PM10 (high vs low exposure) |

Hazard Ratios and 95% confidence intervals (HR [95% CI]) are reported for each model, while association p-value is reported only for the most adjusted model (see below). Model 1: age, sex and education level completed; Model 2: Model 1 + professional exposure to toxic compounds and working class; Model 3: Model 2 + lifestyles + BMI. Both HR associated with unitary increase of PM10 (µg/m<sup>3</sup>) and for participants exposed to high vs low exposure compared to median PM10 levels (11.6 µg/m<sup>3</sup>) are reported. Legend: AD = Alzheimer's Disease/Dementia; PD = and Parkinson's Disease/Parkinsonisms; PM10 = particulate matter with aerodynamic diameter < 10 µm.

**Figure 2.**
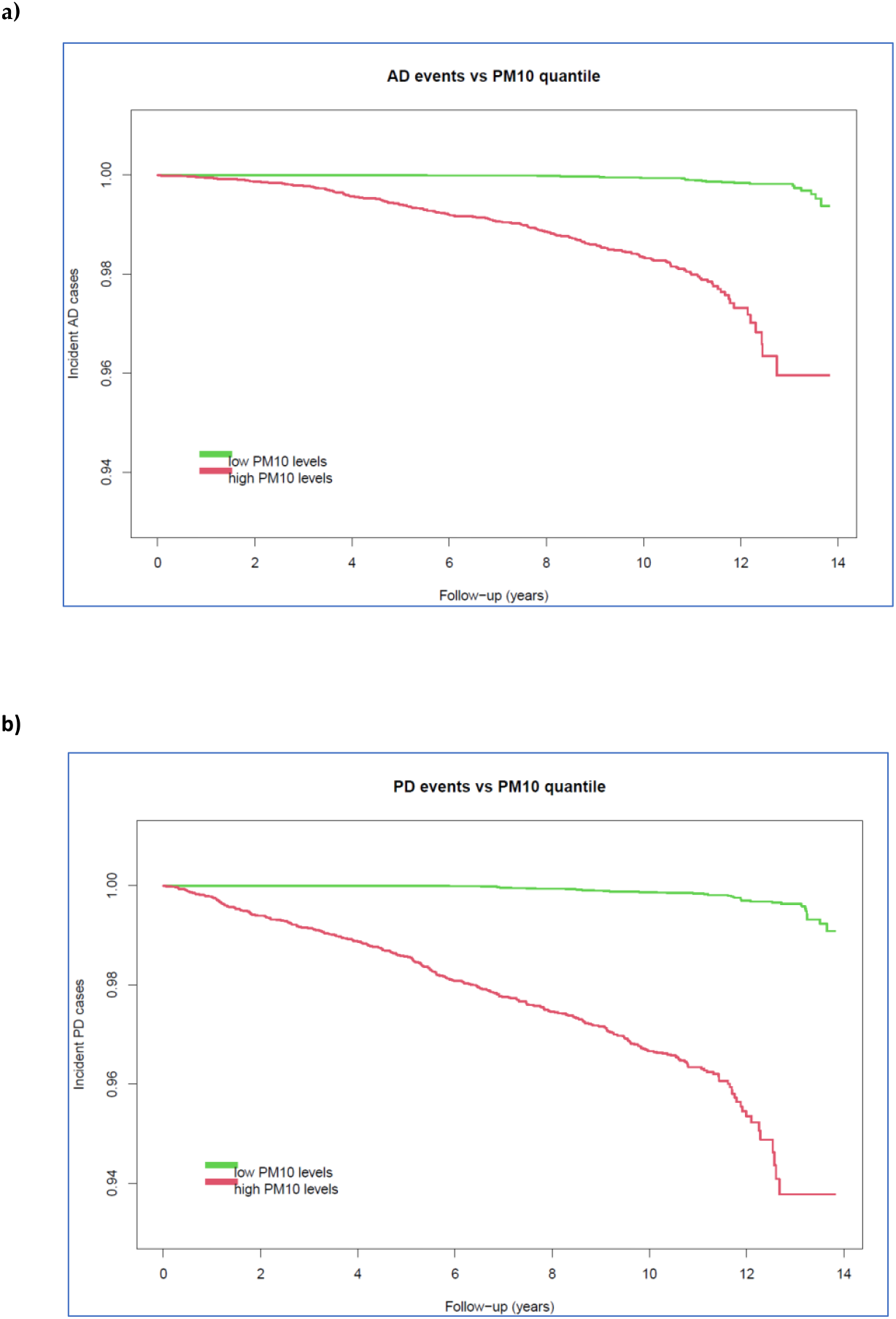
Kaplan Meier curves of incident a) AD and b) PD events vs PM10 quantiles. Incident **a)** dementia and **b)** Parkinson’s disease/parkinsonisms events are compared between the two quantiles of exposure to PM10: below the median (11.6 μg/m3; green) and above the median (red)

When we tested potential roles of inflammatory markers like INFLA-score, GLR, CRP, Plt and WBC, the proportion of the above-mentioned associations explained by these markers was non-significant and negligible (<1%) for both AD and PD (Table 4).

**Table 4.** Proportion of association between air pollution (PM10) and incident AD/PD risk explained by circulating inflammation.

| *Outcome* | *INFLA-score* | *CRP* | *Plt* | *WBC* | *GLR* |
| --- | --- | --- | --- | --- | --- |
|  | *PTE (SD),* | *PTE (SD),* | *PTE (SD),* | *PTE (SD),* | *PTE (SD),* |
|  | *p-value* | *p-value* | *p-value* | *p-value* | *p-value* |
| *AD* | 0.59 (0.53)% | 0.57 (0.49)% | 0.57 (0.52)% | 0.61 (0.55)% | 0.59 (0.55)% |
|  | p = 0.43 | p = 0.42 | p = 0.42 | p = 0.45 | p = 0.42 |
| *PD* | 0.67 (0.61)% | 0.71 (0.64)% | 0.66 (0.65)% | 0.70 (0.60)% | 0.68 (0.63)% |
|  | p = 0.71 | p = 0.74 | p = 0.70 | p = 0.74 | p = 0.72 |

We report percentage of total effect (PTE) explained by the INFLA-score [7] and its component biomarkers in the association between PM10 levels and incident AD and PD risk, along with relevant standard deviation (SD) and empirical p-value, as computed through bootstrapping simulations. These estimates refer to Model 3, adjusted for sociodemographic, professional and lifestyle covariates.
Abbreviations: AD = dementia; PD : Parkinson’s disease/parkinsonisms; CRP = C-reactive protein; Plt = platelets count; GLR = granulocyte-to-lymphocyte ratio; WBC = white blood cells count.

## Discussion

In the present manuscript, we analyzed the relationship between air pollution and incident neurodegenerative risk in an Italian population, identifying a notable influence of PM10 levels on an increased risk of both dementia forms and Parkinson’s disease/parkinsonisms. Interestingly, this influence was independent on diverse other factors representing known risk/protective factors for AD and PD, including lifestyles like physical activity, smoking, drinking and adherence to Mediterranean Diet [35,36], and on professional factors like working class and exposure to toxic compounds. While a link between PM10 and neurodegenerative risk has been long hypothesized, only few studies supported this through statistical evidence – especially for AD – and recent meta-analyses revealed contrasting results and a yet unclear relationship [13,15,16]. Indeed, most of the studies have so far supported an association of increased AD and PD risk with PM2.5 and NO2 [13,15,17,18,37,38], but findings on PM10 are less consistent. A geospatial analysis of PD cases and controls reported a significant difference in the mean annual NO2 and PM10 levels between areas where PD cases were concentrated (hotspots) and those where they were not (coldspots) [23]. These findings are in line with a large nested case-control study from a Chinese health insurance cohort, where PM10 was the only pollutant reported to influence an increase in PD risk, among many others [21]. However, other cohort studies found no evidence of association with PM10 [17,20,39,40], as well as case-control studies [22,41,42], and even meta-analyses [30,43].

Other works identified associations between PM2.5 and clinical events related to these disorders, like an increased risk of hospitalization for dementia causes (e.g [44]) or an increased mortality in Parkinson’s disease patients [16]. Some studies like the Rome Longitudinal Study reported a positive association of both PM2.5 and PM10 with incident hospitalization risk for vascular dementia, but a negative association with Alzheimer dementia [19], and no association with Parkinson’s disease [20]. Other studies have found associations between exposure to air pollutants – particularly PM2.5, PM10 and NO2 - and neuropathological hallmarks of AD and PD [25,45,46,47], neuroimaging endophenotypes like a reduced prefrontal cortex [48], hippocampal volume [49], and cortical thickness in the temporal lobe [50].

In spite of these promising findings, the molecular mechanism of action of these pollutants remains largely unclear. PM represent one of the main suspect to play a role in the pathophysiology of neurodegeneration since they are small enough to enter the lungs and spread through the brain, possibly through alteration of the blood brain barrier, oxidative stress, microglia activation and brain inflammation [8,31,47,51], but also through the alteration of connectivity among different brain regions [13]. In spite of the above mentioned role of neuroinflammation in both AD and PD pathophysiology, and of experimental evidence suggesting that circulating inflammation may somehow mediate the effect of pollution on neurodegenerative risk [6,24,25], our analysis did not reveal any significant explanatory effect by these markers, suggesting that other mediating pathways may act in this link. Among them, biological aging, the actual underlying age of an organism, which can be measured through different algorithms and biomedical data sources (e.g. epigenetic variations, blood markers, brain imaging) may represent a key player. Indeed, both long and short term exposure to air pollution was already reported to influence telomere attrition and epigenetic aging [52,53,54,55], which very well predict incident risks of dementia and Parkinson’s disease [56,57]. This represents a promising hypothesis to investigate in the future, along with gene-by-environment and environment-by-environment interactions [13,58], which show the potential to further clarify how pollution and other exposome layers may influence incident neurodegenerative risk.

Of note, one of the pollutants most reported to have an influence on AD and PD risk by previous studies, NO2 [13], did not reveal any significant association in the present work. While this may depend on the different approach used here to investigate air pollution exposure while controlling for collider bias – namely through PC scores rather than single pollutant levels – this finding is not that surprising in view of the inconsistent results published so far for this and other compounds, which may be due to the same elements of heterogeneity across studies mentioned above for PM10.

### Strengths and Limitations

The present work presents strengths like the comprehensive approach to multiple air pollutants from diverse sources and the simultaneous assessment of many lifestyles and professional factors which may influence incident neurodegenerative risk, which allowed us to identify a clear influence of PM10 on AD and PD, independent on all other risk and protective factors tested. Moreover, although a role of inflammation has been often hypothesized, we are not aware of any study testing potential explanations of the link between pollution and neurodegenerative disorders through a diverse set of circulating inflammation markers, tagging cytokine-related, hemostatic and cellular/immune components of inflammation. However, our work also has some limitations. First, the use of land measurements to interpolate exposure maps may not be as precise as maps integrating land and satellite data, as well as the fact that data for the first year of recruitment (2005) were not available in the ARPA Molise database. However, Molise is a rather rural region which does never experience sudden massive changes in the degree of anthropic activities, hence the levels of air pollutants tend to remain stable across the years. This, along with the dimensionality reduction approach used, which was necessary to reduce collinearity biases, may explain the large effect sizes observed. We plan to further investigate these associations using environmental data from other independent European sources, which will be made available in the future. Third, the algorithm used for defining incident neurodegenerative cases may have led to classify as AD/PD cases subjects affected by related disorders or with a yet unclear diagnosis, a problem often affecting neurological clinical practice. However, we remained prudent in the definition of the neurodegenerative disorders analyzed and still this does not affect the focus of the manuscript, on the relationship between pollution and incident neurodegenerative risk in the population, which needs modifiable risk factors to be identified, whatever the clinical diagnosis. Fourth, circulating inflammation does not necessarily reflect neuroinflammation, which may explain the low proportions of associations explained by circulating inflammatory markers. Last, the simultaneous assessment of environmental exposures and circulating inflammation markers does not allow us to infer direction of causality between them, hence we could not perform a formal mediation analysis. We are now working on improving many of these aspects, so to run more powerful analyses with more precise exposure estimates and longer and validated neurological follow-up.

## Data Availability

All data produced in the present study are available upon reasonable request to the authors

## Authors’ contributions

AG, SC, LI, SM and FG, MBD, GdG, CC, GV and MMF contributed to the concept and design of the work and/or to the interpretation of data. SC, TP, MR and SF managed data collection, curation and elaboration. AC, FM and GZ carried out elaboration, quality control and curation of environmental data. AG, SC and AT performed data analysis. AG, AT, AC, FM and GZ wrote a first draft of the manuscript, with critical contributions from all the co-authors. All the co-authors approved the final version of the manuscript.

## Funding

The enrollment phase of the Moli-sani Study was supported by unrestricted research grants from the Pfizer Foundation (Rome, Italy), the Italian Ministry of University and Research (MIUR, Rome, Italy)—Programma Triennale di Ricerca, Decreto no.1588, and Instrumentation Laboratory, Milan, Italy. The follow-up phase of the Moli-sani Study (assessment of incident cases) was partially supported the Italian Ministry of Health (PI GdG, CoPI SC; grant no. RF-2018-12367074). The collection and elaboration of environmental data was supported by the Italian Ministry of Economic Development (PLATONE project, bando “Agenda Digitale” PON I&C 2014-2020; Prog. n. F/080032/01-03/X35). The present analyses were partially supported by the INAIL-Bric 2019 ID 47 project (Italian National Institute for Insurance against Accidents at Work; call 2019, project CUP code: F24I19000630008).

No funder had a role in study design, collection, analysis, interpretation of data, writing of the manuscript, and decision to submit this article for publication.

## Acknowledgements

The Moli-sani research group thanks the Associazione Cuore Sano Onlus (Campobasso, Italy) for its cultural support, Marno Srl (Rosignano Marittimo, LI) and Innovation Group Scrl (Sant’Agapito, IS) for the record linkage with electronic health data.

## Data Availability Statement

The data underlying this article will be shared upon reasonable request to the corresponding author. The data are stored in an institutional repository (https://repository.neuromed.it) and access is restricted by the ethics approval and the legislation of the European Union.

## URLs

ARPA Molise: https://www.arpamolise.it/

Kriging algorithm: https://desktop.arcgis.com/en/arcmap/10.3/tools/3d-analyst-toolbox/how-kriging-works.htm

GeoKettle: http://www.geokettle.org/

HERE: https://www.here.com/

QGis: https://www.qgis.org/en/site/

R: https://www.r-project.org/

## Notes

**Conflict of interest** Nothing to declare.

### Competing Interest Statement

The authors have declared no competing interest.

### Author Declarations

The Moli-sani Study was approved by the ethical committee of the Catholic University of Rome (approval nr: P99, A-931/03-138-04/CE/2004, 11 February 2004) and all the participants provided written informed consent.

