## Supplemental_material for "Prominent role of PM10 but not of circulating inflammation in the link between air pollution and the risk of neurodegenerative disorders"

### Supplementary Materials

#### *Circulating biomarkers*

Blood samples were obtained between 7 and 9 AM from participants who had fasted overnight and had refrained from smoking for at least 6 hours (h). Biochemical analyses were performed in the centralised Moli-sani laboratory, as described elsewhere[1]. Haemochromocytometric analyses were performed by cell counter (Coulter HMX, Beckman Coulter, Milan, Italy) within 3 h from venepuncture. All haemochromocytometric analyses (white blood cell and platelet counts, granulocyte % and lymphocyte %) were performed by cell counter (Coulter HMX, Beckman Coulter, Milan, Italy) within 3 h from venipuncture. High sensitivity C reactive protein (CRP) was measured in fresh serum, by a latex particle-enhanced immunoturbidimetric assay (ILab 350 Instrumentation Laboratory, Milan, Italy).

#### *Definition of covariates*

*Smoking status* was defined as a three-class variable based on cigarette smoking habits of participants: *smokers, ex-smokers* (i.e. subjects who quitted at least one year before the interview) and *never-smokers* (reference class).

Leisure-time *physical activity* was assessed through a structured questionnaire and expressed as daily energy expenditure in metabolic equivalent task-hours (MET-h/day)[2].

Food intake was assessed through the validated Italian EPIC food frequency questionnaire [3]. Adherence to *Mediterranean Diet* was defined according to the Mediterranean Diet Score, ranging from 0 (low adherence) to 9 (high adherence)[4]. Then we defined three adherence classes: low (Trichopoulou score 0-3, used as reference), moderate (4-6) and high (7-9). The EPIC questionnaire also allowed to compute *alcohol consumption* habits along with additional questions, as described by [5]. Participants were classified based on the estimated daily alcohol intake in the year before enrolment: life-time abstainers, former drinkers, occasional drinkers and current drinkers who drank 1–12 (reference class), 12.1–24, 24.1–48 and >48 g/day.

Height and weight were measured for each participant, as well as waist circumference (cm), which was measured in the middle between the 12$^{th}$ rib and the iliac crest, while hip circumference (cm) was measured around the buttocks. Then *Body mass index (BMI)* was calculated, and participants were grouped into three categories: <25, 25–29.9, and ≥30 kg/m$^2$.

Socioeconomic status (SES) information was self-reported and/or collected by a structured questionnaire administered by trained personnel. *Educational attainment* was defined as the education level completed and subjects were divided into four classes: None/Primary (reference), Lower secondary, Upper secondary and Post-secondar*y*. Current *occupational social class* was classified based on the Registrar General's occupation classification scheme, and ranked as previously described by [6]. Based on this rank, five occupational classes were defined: professional/managerial, skilled non-manual, skilled manual, partly skilled/unskilled and unemployed/unclassified (reference).

Professional exposure to potentially toxic compounds, like concrete, eternit, aluminum, starchy and paper-derived compounds was also self-reported by each participant (yes/no).

Prevalent *diabetes* and *hyperlipidemia* were defined as dichotomous variables (Yes/No), based on the reported and verified use of specific drugs for the treatment of these disorders. Prevalent *cardiovascular disease (CVD)* and *cancer* were defined as binary variables, classified into subjects reporting medical history of the disease (possibly supported by medical documentation or by the use of specific drugs), and those with no medical history of the disease.

**Figure S1.** Scree plot of the first 20 principal components extracted from air pollution data.

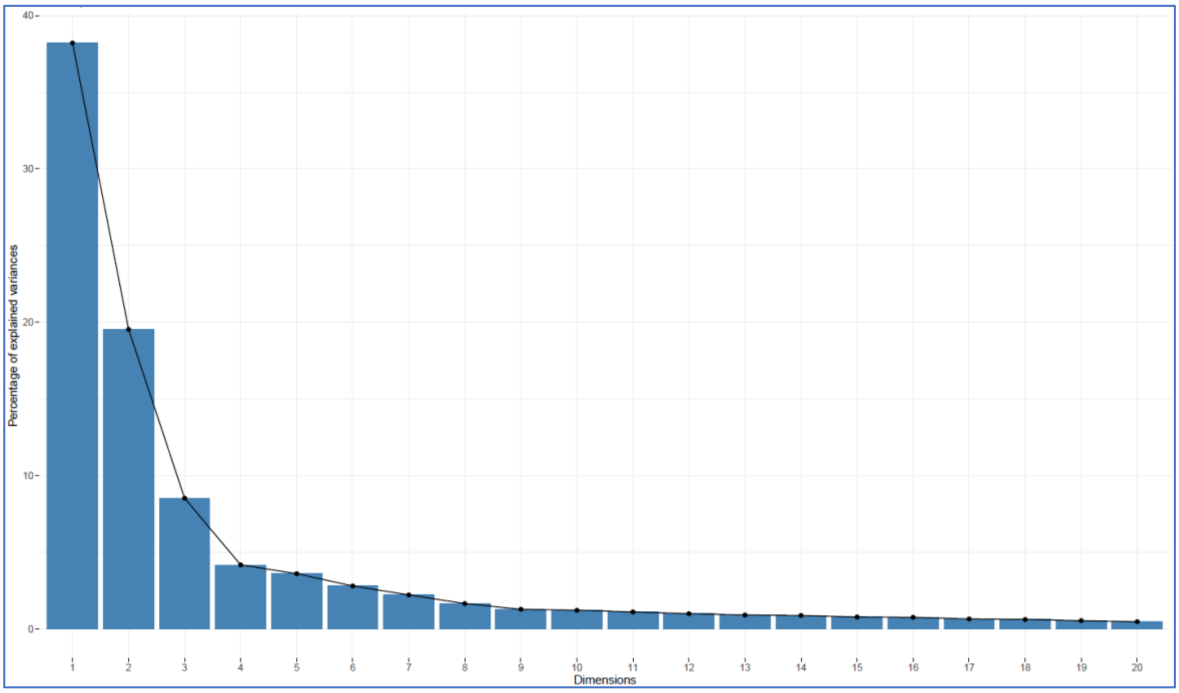


Percentage of variance shared across all the yearly air pollution levels (for ten pollutants and 13 years, 2006-2018) is reported for the first 20 principal components extracted.

**Figure S2.** Biplots of the first three of principal components extracted from air pollution data.

a)

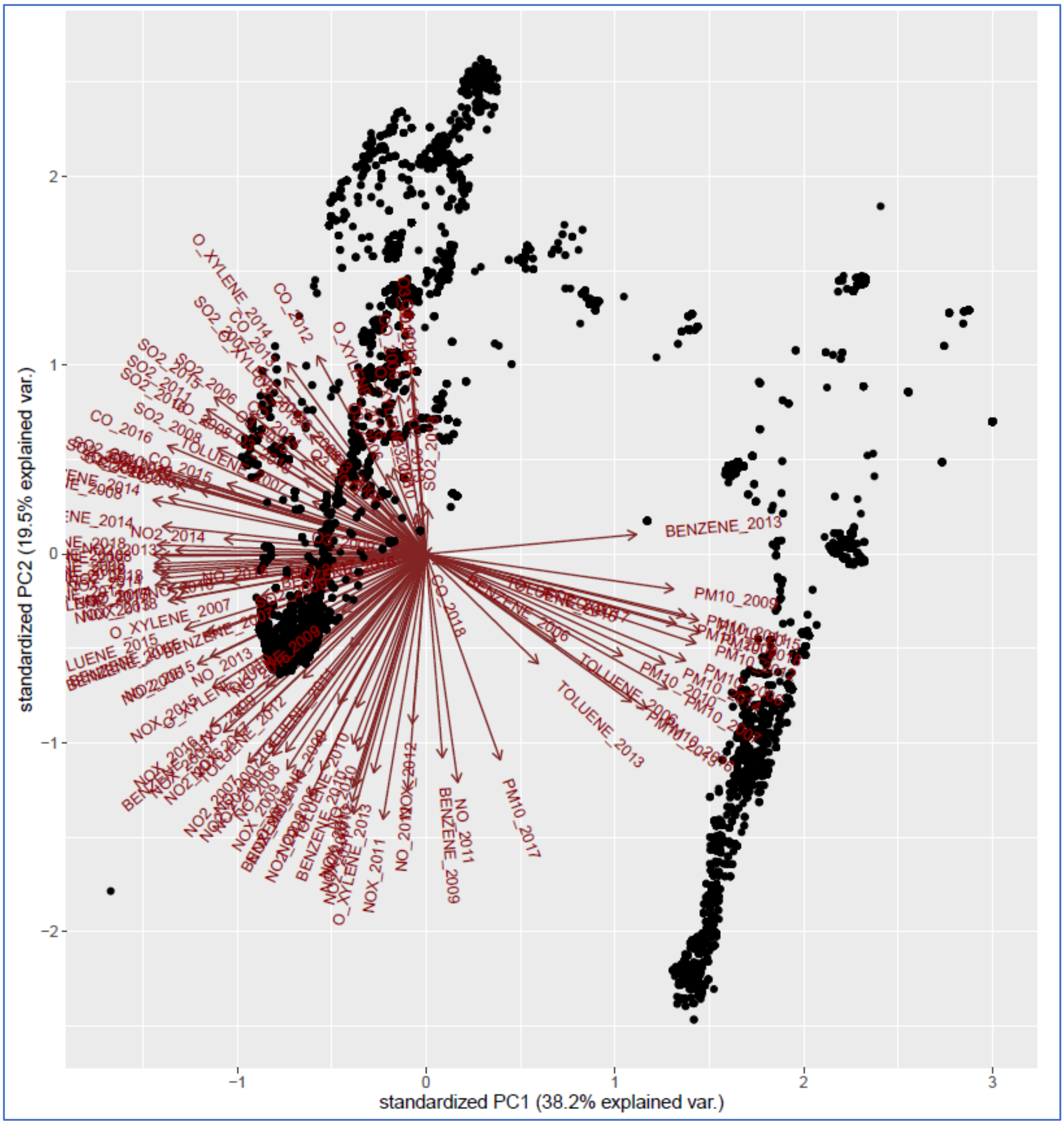

b)

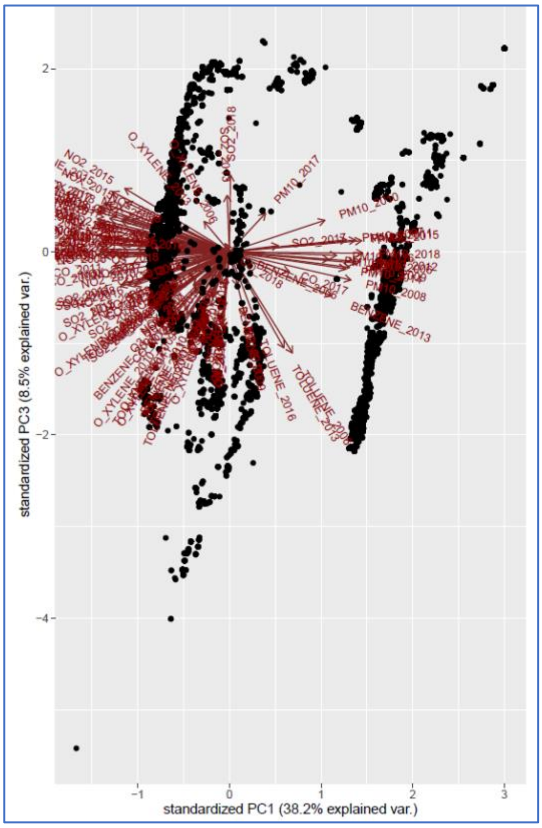


c)

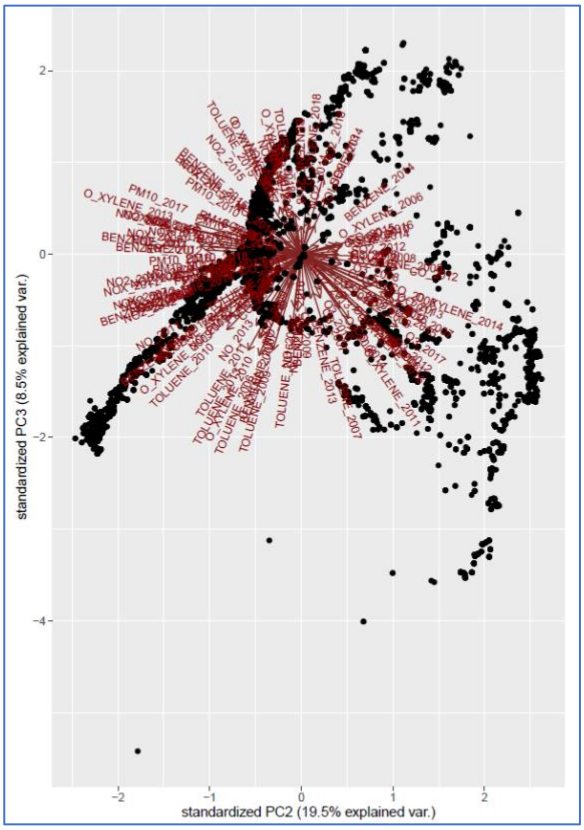


Each dot represents a participant, while arrows represent the loadings of each yearly air pollution exposure on the PC scores plotted.

**Figure S3.** Spearman correlation plots of average pollutants levels for the analysis of incident risk of a) AD and b) PD.

**a)**

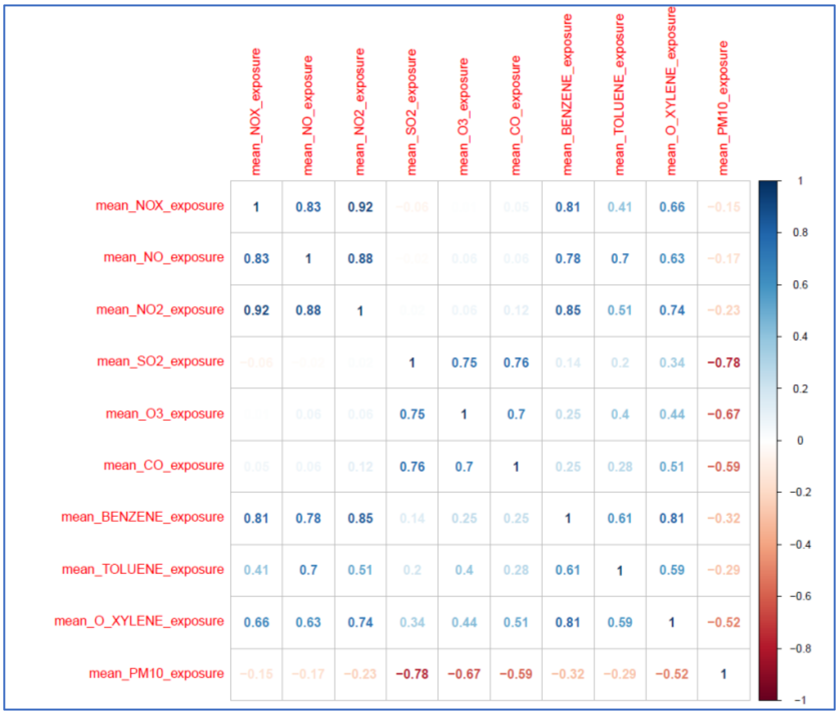


b)

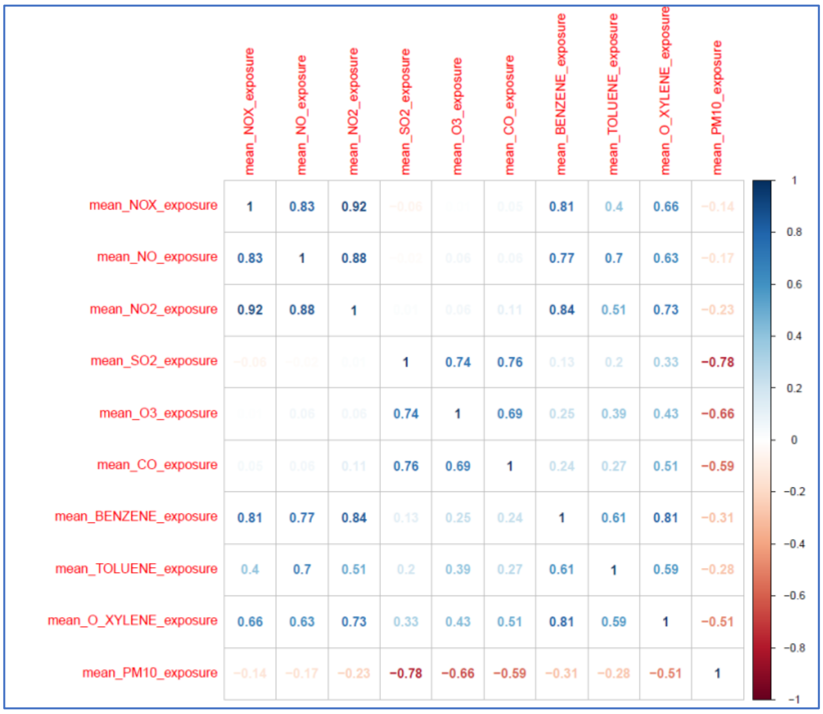

**Figure S4.** Distribution of PM10 exposure levels averaged over the follow-up time in the Moli-sani cohort.

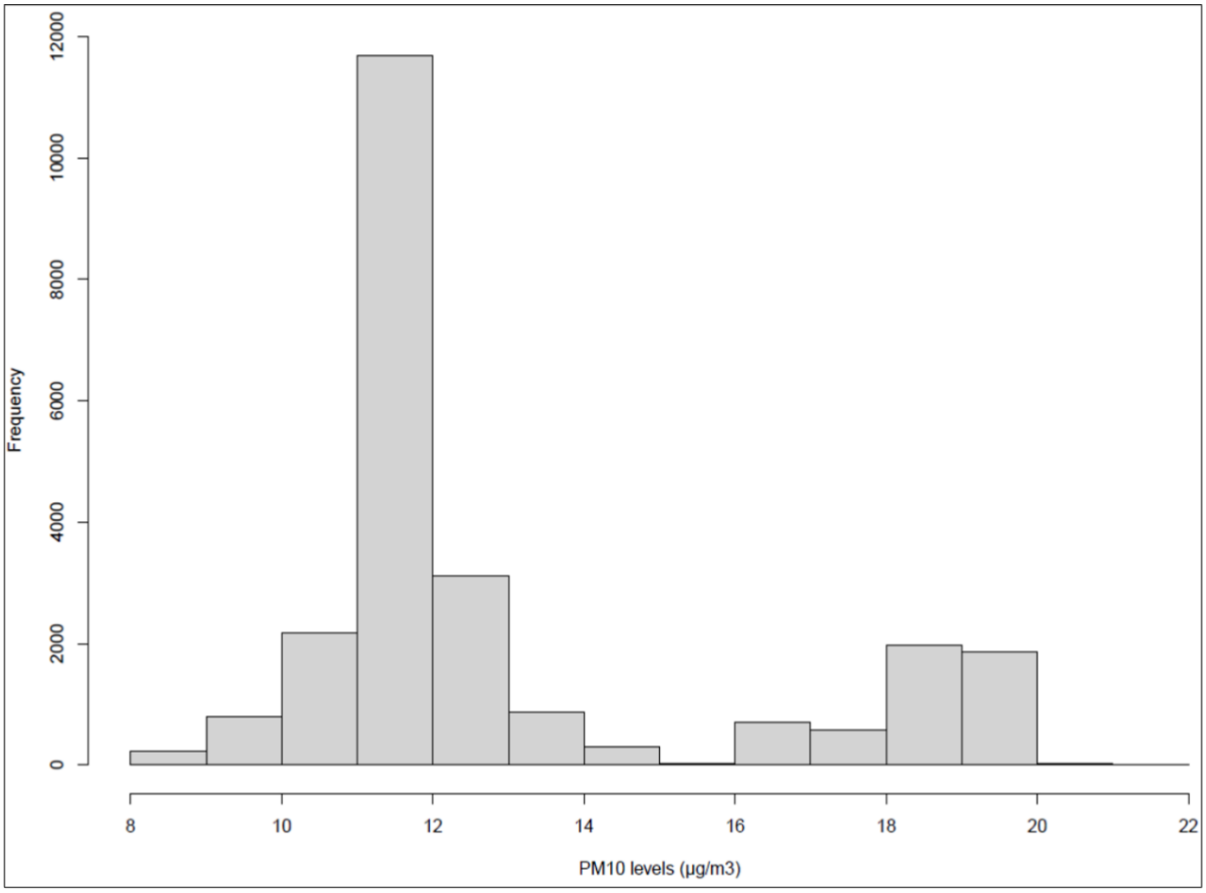


Median (IQR): 11.64 (1.61) μg/m3.

**Table S1.** Definition of incident Alzheimer/dementia and Parkinson/parkinsonisms events in the Moli-sani cohort.

| Disease | Parkinson/Parkinsonisms | Alzheimer/dementia |
|---|---|---|
| **Regional drug prescription registry** | | |
| ATC code | N04: Antiparkinsonian drugs | N06D: Anti-dementia drugs |
| **Hospital discharge records (diagnosis)** | | |
| ICD-9 cm | 332: Parkinson's disease<br>332.0: Parkinsonism or other Parkinson's disease<br>332.1: Secondary Parkinsonism | 331.0: Alzheimer's disease<br>331.X: Other neurodegenerative processes |
| **National (ReNCaM) mortality registry (cause of death)** | | |
| ICD-9 cm | 332: Parkinson's disease | 331: Alzheimer's disease |

**Table S2.** Full results of the association analysis of incident a) AD and b) PD risk.

**a)**

| Variable | HR [95% CI] | Z-score | P |
|---|---|---|---|
| **Age (years)** | **1.13 [1.11-1.15]** | **14.7** | **1.00E-47** |
| **air pollution PC1** | **1.06 [1.04-1.08]** | **9.12** | **8.65E-08** |
| air pollution PC2 | 0.96 [0.93-0.99] | 1.92 | 1.41E-02 |
| high BMI (≥30 kg/m2) vs low (<25 kg/m2) | 0.69 [0.45-1.06] | -1.74 | 9.37E-02 |
| professional exposure to concrete compounds | 1.5 [0.93-2.43] | -1.61 | 9.87E-02 |
| professional exposure to starchy compounds | 1.45 [0.91-2.31] | -1.59 | 1.20E-01 |
| drinker (moderate vs mild-occasional) | 0.67 [0.41-1.11] | 1.5 | 1.20E-01 |
| drinker (previous vs mild-occasional) | 0.5 [0.2-1.27] | -1.25 | 1.45E-01 |
| professional exposure to aluminium compounds | 0.56 [0.23-1.35] | 1.22 | 1.98E-01 |
| Mediterranean Diet (high vs low adherence) | 1.31 [0.77-2.25] | -1.08 | 3.19E-01 |
| drinker (heavy vs mild-occasional) | 0.79 [0.49-1.28] | 1.01 | 3.38E-01 |
| professional exposure to paper-derived compounds | 1.43 [0.62-3.31] | -0.92 | 3.99E-01 |
| education (post-secondary vs none/primary) | 0.73 [0.35-1.53] | -0.72 | 4.10E-01 |
| Leisure time physical activity (MET-h/day) | 0.98 [0.95-1.02] | 0.59 | 4.19E-01 |
| smoking (previous vs non-smoking) | 1.12 [0.77-1.63] | -0.56 | 5.63E-01 |
| Mediterranean Diet (moderate vs low adherence) | 1.1 [0.78-1.55] | 0.49 | 5.93E-01 |
| drinker (very heavy vs mild-occasional) | 0.86 [0.46-1.59] | -0.43 | 6.23E-01 |
| smoking (current vs non-smoking) | 0.88 [0.52-1.49] | -0.31 | 6.45E-01 |
| drinker (never vs mild-occasional) | 1.09 [0.72-1.65] | -0.22 | 6.79E-01 |
| moderate BMI (25-29.9 kg/m2) vs low (<25 kg/m2) | 1.05 [0.72-1.53] | -0.21 | 8.00E-01 |
| Sex (male vs female) | 0.95 [0.62-1.44] | 0.18 | 8.01E-01 |
| education (lower secondary vs none/primary) | 0.95 [0.6-1.51] | -0.08 | 8.24E-01 |
| education (upper secondary vs none/primary) | 0.95 [0.58-1.55] | 0.07 | 8.37E-01 |
| air pollution PC3 | 1 [0.96-1.05] | 0.01 | 8.66E-01 |
| professional exposure to eternit compounds | 0.91 [0.22-3.81] | 0.01 | 8.95E-01 |
| occupational class (professional/managerial vs unemployed/unclassified) | [0; Inf] | 0.01 | 9.92E-01 |
| occupational class (non-manual worker vs unemployed/unclassified) | [0; Inf] | 0.01 | 9.92E-01 |
| occupational class (manual skilled worker vs unemployed/unclassified) | [0; Inf] | 0.01 | 9.92E-01 |
| occupational class (retired/housewife vs unemployed/unclassified) | [0; Inf] | 0.00 | 9.92E-01 |
| occupational class (manual unskilled worker vs unemployed/unclassified) | [0; Inf] | 0.00 | 9.92E-01 |

**b)**

| Variable | HR [95% CI] | Z-score | P |
|---|---|---|---|
| **Age (years)** | **1.08 [1.07-1.09]** | **14.7** | **2.03E-39** |
| **air pollution PC1** | **1.05 [1.03-1.06]** | **9.12** | **6.56E-09** |
| Sex (male vs female) | 1.45 [1.08-1.93] | 1.92 | 1.20E-02 |
| drinker (very heavy vs mild-occasional) | 0.66 [0.42-1.03] | -1.74 | 6.72E-02 |
| drinker (heavy vs mild-occasional) | 0.73 [0.51-1.03] | -1.61 | 7.52E-02 |
| professional exposure to paper-derived compounds | 1.62 [0.95-2.76] | -1.59 | 7.88E-02 |
| drinker (moderate vs mild-occasional) | 0.74 [0.52-1.06] | 1.5 | 1.04E-01 |
| professional exposure to eternit compounds | 0.36 [0.09-1.48] | -1.25 | 1.57E-01 |
| moderate BMI (25-29.9 kg/m2) vs low (<25 kg/m2) | 0.83 [0.62-1.1] | 1.22 | 1.85E-01 |
| professional exposure to aluminium compounds | 0.69 [0.38-1.27] | -1.08 | 2.32E-01 |
| occupational class (manual skilled worker vs unemployed/unclassified) | 3.25 [0.45-23.53] | 1.01 | 2.44E-01 |
| occupational class (manual unskilled worker vs unemployed/unclassified) | 2.89 [0.4-20.99] | -0.92 | 2.94E-01 |
| professional exposure to concrete compounds | 1.2 [0.84-1.71] | -0.72 | 3.09E-01 |
| occupational class (non-manual worker vs unemployed/unclassified) | 2.58 [0.36-18.62] | 0.59 | 3.47E-01 |
| occupational class (professional/managerial vs unemployed/unclassified) | 2.55 [0.35-18.56] | -0.56 | 3.56E-01 |
| drinker (previous vs mild-occasional) | 1.24 [0.77-1.99] | 0.49 | 3.81E-01 |
| Leisure time physical activity (MET-h/day) | 0.99 [0.96-1.02] | -0.43 | 3.99E-01 |
| occupational class (retired/housewife vs unemployed/unclassified) | 2.28 [0.3-17.15] | -0.31 | 4.22E-01 |
| education (lower secondary vs none/primary) | 0.88 [0.63-1.23] | -0.22 | 4.42E-01 |
| smoking (previous vs non-smoking) | 0.9 [0.69-1.19] | -0.21 | 4.73E-01 |
| air pollution PC2 | 1.01 [0.99-1.03] | 0.18 | 5.44E-01 |
| education (post-secondary vs none/primary) | 1.14 [0.69-1.89] | -0.08 | 5.97E-01 |
| smoking (current vs non-smoking) | 0.92 [0.66-1.28] | 0.07 | 6.18E-01 |
| air pollution PC3 | 0.99 [0.96-1.02] | 0.01 | 6.30E-01 |
| Mediterranean Diet (high vs low adherence) | 1.08 [0.73-1.6] | 0.01 | 6.93E-01 |
| professional exposure to starchy compounds | 1.07 [0.75-1.53] | 0.01 | 7.13E-01 |
| high BMI (≥30 kg/m2) vs low (<25 kg/m2) | 0.96 [0.72-1.29] | 0.01 | 8.01E-01 |
| drinker (never vs mild-occasional) | 0.97 [0.71-1.32] | 0.01 | 8.42E-01 |
| Mediterranean Diet (moderate vs low adherence) | 0.98 [0.76-1.25] | 0.00 | 8.57E-01 |
| education (upper secondary vs none/primary) | 1 [0.69-1.44] | 0.00 | 9.82E-01 |

Hazard Ratios and 95% confidence intervals (HR [95% CI]) are reported for all the covariates tested in the most adjusted model (Model 3: age + sex + education level completed + professional exposure to toxic compounds + working class + lifestyles + BMI). Associations surviving Bonferroni correction for multiple testing (p-value < 0.008) are highlighted in bold.

**Table S3.** Results of the sensitivity analysis of incident AD and PD risk vs PM10 levels, after removal of early onset cases.

| Outcome | Exposure | Model 3: HR [95% CI], p |
|---|---|---|
| AD | PM10 ($\mu g/m^3$) | 1.25 [1.19-1.31], $4.9\times10^{-19}$ |
| AD | PM10 (high vs low exposure) | 20.76 [11.67-36.96], $6.3\times10^{-25}$ |
| **PD** | PM10 ($\mu g/m^3$) | 1.19 [1.15-1.24], $7.3\times10^{-22}$ |
| **PD** | PM10 (high vs low exposure) | 13.10 [9.08-18.89], $4.2\times10^{-43}$ |

Hazard Ratios and 95% confidence intervals (HR [95% CI]) and p-values are reported for the most adjusted model (Model 3: age + sex + education level completed + professional exposure to toxic compounds + working class + lifestyles + BMI). Both HR associated with unitary increase of PM10 ($\mu g/m^3$) and for participants exposed to high vs low exposure compared to median PM10 levels (11.6 $\mu g/m^3$) are reported. Legend: AD = Alzheimer's Disease/Dementia; PD = and Parkinson's Disease/Parkinsonisms; PM10 = particulate matter with aerodynamic diameter < 10 μm.

**Moli-sani Study Investigators**

The enrolment phase of the Moli-sani Study was conducted at the Research Laboratories of the Catholic University in Campobasso (Italy), the follow up of the Moli-sani cohort is being conducted at the Department of Epidemiology and Prevention of the IRCCS Neuromed, Pozzilli, Italy.

**Steering Committee:** Licia Iacoviello*°(Chairperson), Giovanni de Gaetano* and Maria Benedetta Donati*.

**Scientific Secretariat:** Marialaura Bonaccio*, Americo Bonanni*, Chiara Cerletti*, Simona Costanzo*, Amalia De Curtis*, Augusto Di Castelnuovo§, Alessandro Gialluisi*°, Francesco Gianfagna°§, Mariarosaria Persichillo*, Teresa Di Prospero* (Secretary).

**Safety and Ethical Committee:** Jos Vermylen (Catholic University, Leuven, Belgio) (Chairperson), Renzo Pegoraro (Pontificia Accademia per la Vita, Roma, Italy), Antonio Spagnolo (Catholic University, Roma, Italy).

**External Event Adjudicating Committee**: Deodato Assanelli (Brescia, Italy), Livia Rago (Campobasso, Italy).

**Baseline and Follow-up Data Management:** Simona Costanzo* (Coordinator), Marco Olivieri (Campobasso, Italy), Teresa Panzera*.

**Data Analysis:** Augusto Di Castelnuovo§ (Coordinator), Marialaura Bonaccio*, Simona Costanzo*, Simona Esposito*, Alessandro Gialluisi*°, Francesco Gianfagna°§, Sabatino Orlandi*, Emilia Ruggiero*, Alfonsina Tirozzi*.

**Biobank, Molecular and Genetic Laboratory:** Amalia De Curtis* (Coordinator), Sara Magnacca§, Fabrizia Noro*, Alfonsina Tirozzi*.

**Recruitment Staff:** Mariarosaria Persichillo* (Coordinator), Francesca Bracone*, Teresa Panzera*.

**Communication and Press Office:** Americo Bonanni*.

**Regional Institutions:** Direzione Generale per la Salute - Regione Molise; Azienda Sanitaria Regionale del Molise (ASReM, Italy); Agenzia Regionale per la Protezione Ambientale del

Molise (ARPA Molise, Italy); Molise Dati Spa (Campobasso, Italy); Offices of vital statistics of the Molise region.

**Hospitals:** Presidi Ospedalieri ASReM: Ospedale A. Cardarelli – Campobasso, Ospedale F. Veneziale – Isernia, Ospedale San Timoteo - Termoli (CB), Ospedale Ss. Rosario - Venafro (IS), Ospedale Vietri – Larino (CB), Ospedale San Francesco Caracciolo - Agnone (IS); Casa di Cura Villa Maria - Campobasso; Ospedale Gemelli Molise - Campobasso; IRCCS Neuromed - Pozzilli (IS).

*Department of Epidemiology and Prevention, IRCCS Neuromed, Pozzilli, Italy

°Department of Medicine and Surgery, University of Insubria, Varese, Italy

§Mediterranea Cardiocentro, Napoli, Italy

*Moli-sani Study Past Investigators are available at* [*https://www.moli-sani.org/?page_id=173*](https://www.moli-sani.org/?page_id=173)
